# Vaccine-Elicited Antibody Responses to Influenza H3N2 Subclade K

**DOI:** 10.64898/2026.02.02.26345378

**Authors:** Liping Wang, Samuel J. Nangle, Alejandra Waller-Pulido, Katherine McMahan, Juliana Pereira, Jayeshbhai Chaudhari, Leland O. Barrett, Ritobhas Bhowmik, Keith Ferrugia, Reima Ramsamooj, Emilia Mia Sordillo, Viviana Simon, Harm van Bakel, Heba Mostafa, Andrew Pekosz, Ai-ris Y. Collier, Dan H. Barouch

**Affiliations:** Beth Israel Deaconess Medical Center, Harvard Medical School, Boston, MA, USA; Icahn School of Medicine at Mount Sinai, New York, NY, USA; Johns Hopkins School of Medicine, Baltimore, Maryland, USA; Johns Hopkins Bloomberg School of Public Health, Baltimore, Maryland, USA

**Author notes:** Corresponding author: Dan H. Barouch, M.D., Ph.D., Center for Virology and Vaccine Research, 330 Brookline Avenue, E/CLS-1043, Boston, MA 02115.

## Abstract

Influenza H3N2 subclade K (J.2.4.1) is a genetic branch of H3N2 with 11 mutations in hemagglutinin and currently represents the dominant circulating influenza strain. We evaluated antibody responses to H3N2 subclade K before and after influenza vaccination in 46 healthy individuals. Our data show that baseline antibody responses to two H3N2 subclade K variants were lower than to other H1N1 and H3N2 strains and that antibody responses following vaccination were also less robust to the H3N2 subclade K variants. These data indicate that the H3N2 subclade K strain partially evades both prior H3N2 immunity and the current inactivated influenza vaccine.

---

Influenza H3N2 subclade K (J.2.4.1) is a genetic branch of H3N2 with 11 mutations in hemagglutinin (HA) compared with the A/H3N2/Croatia/10136RV/2023 vaccine strain, of which 8 mutations are on the surface of the HA head (**Figs. 1A, S1**)^1^. Influenza A viruses currently represent approximately 97% of circulating influenza strains, with H3N2 accounting for 87% of influenza isolates and subclade K comprising 89% of H3N2 isolates^2,3^. The rapid expansion of H3N2 subclade K therefore represents a major public health concern^4,5^. Here we report antibody responses to H3N2 subclade K before and after influenza vaccination.

**Figure 1.**
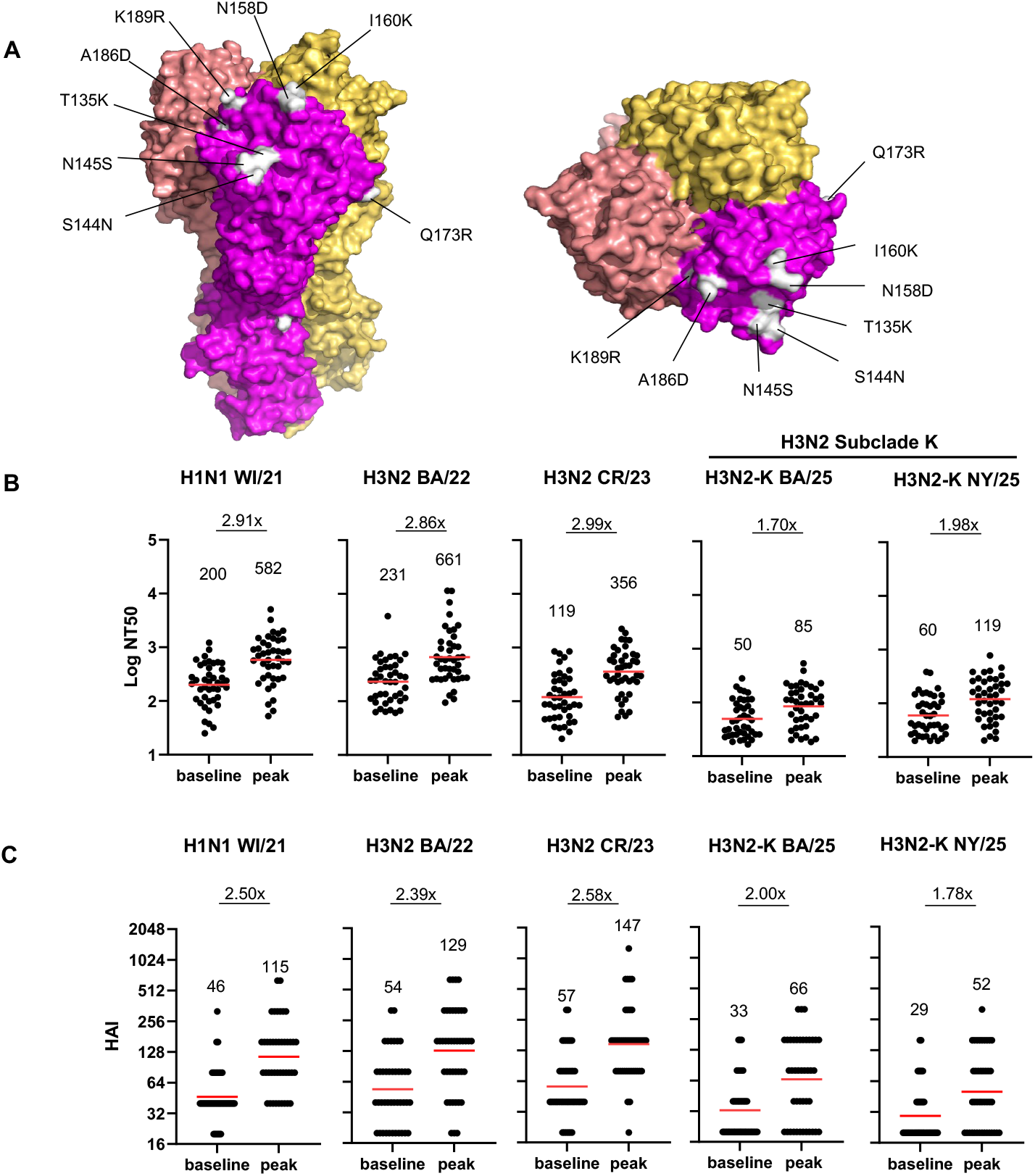
Antibody responses against H1N1 and H3N2 variants following influenza vaccination. (A) Amino acid mutations on the HA head surface of the H3N2 subclade K strain mapped onto the crystal structure of HA trimer from the H3N2 A/Croatia/10136RV/2023 vaccine strain (PDB_ 9ZRU). Each monomer is shown in a distinct color, and mutations on one monomer are shown in white and labeled. Structural visualization was performed using the PyMOL Molecular Graphics System. (B) Neutralizing antibody titers (NT50) are shown before (baseline) and 3 weeks after (peak) trivalent inactivated influenza vaccination to A/H1N1/Wisconsin/67/2021 (H1N1 WI/21), A/H3N2/Baltimore/JH-335/2022 (H3N2 BA/22), A/H3N2/Croatia/10136RV/2023 (H3N2 CR/23), and two H3N2 subclade K strains A/H3N2-K/Baltimore/JH-1365/2025 (H3N2-K BA/25) and A/H3N2-K/New York City/PX23710/2025 (H3N2-K NY/25). (C) HAI titers are shown before (baseline) and 3 weeks after (peak) vaccination. In (B) and (C), red bars indicate geometric mean titers (GMTs), which are displayed numerically above each group. Fold increases from baseline to peak are also shown above each panel.

We assessed influenza-specific antibody responses in 46 healthy adults who received the trivalent inactivated influenza vaccine in fall 2025 in Boston, MA **(Table 1)**. Serum samples were collected prior to immunization (baseline) and at week 3 following immunization (peak). Neutralizing antibody (NAb) titers were measured against the current vaccine strains A/H1N1/Wisconsin/67/2021 (H1N1 WI/21) and A/H3N2/Croatia/10136RV/2023 (H3N2 CR/23), a prior circulating H3N2 strain A/H3N2/Baltimore/JH-335/2022 (H3N2 BA/22), and two H3N2 subclade K strains A/H3N2-K/Baltimore/JH-1365/2025 (H3N2-K BA/25) and A/H3N2-K/New York City/PX23710/2025 (H3N2-K NY/25). The two H3N2 subclade K strains have identical amino acid sequences in HA but differ by five amino acids in internal proteins (NP, NS1, PB1, PA).

**Table 1.**
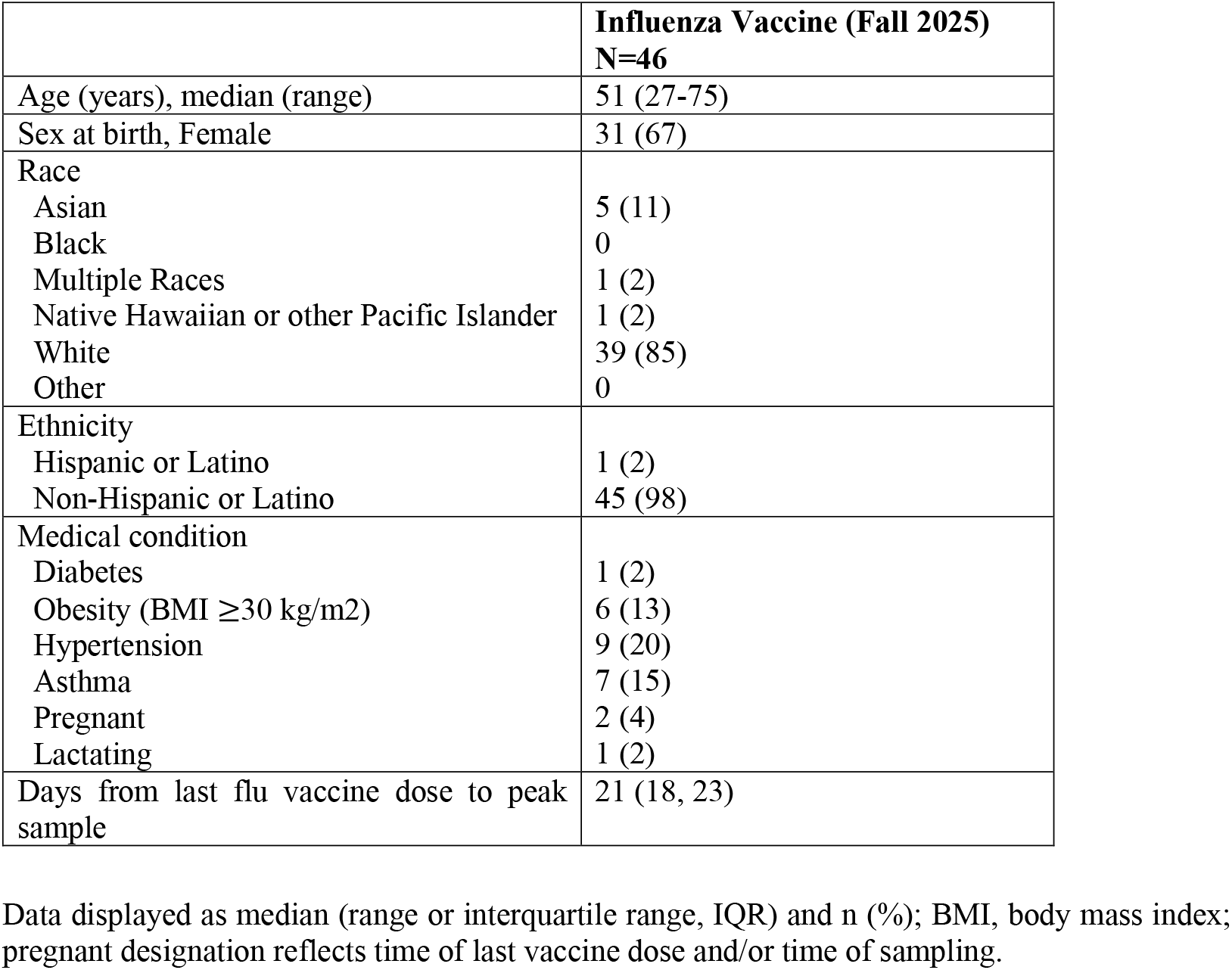
Study population.

NAb geometric mean titers (GMTs) against H1N1 WI/21, H3N2 BA/22, H3N2 CR/23, H3N2-K BA/25, and H3N2-K NY/25 were 200, 231, 119, 50, and 60 at baseline and increased to 582, 661, 356, 85, and 119 at peak immunogenicity, respectively (**Fig. 1B**), reflecting a 2.86- to 2.99-fold increase in NAb titers against the prior H1N1 and H3N2 strains but a lower 1.70- to 1.98-fold increase in NAb titers against the H3N2-K strains. Baseline responses to the H3N2-K strains were 2.0- to 4.6-fold lower than to the prior H1N1 and H3N2 strains, and peak responses to the H3N2-K strains following vaccination were 3.0- to 7.8-fold lower than to the prior H1N1 and H3N2 strains.

Hemagglutination inhibition (HAI) titers showed a similar pattern. HAI GMTs against H1N1 WI/21, H3N2 BA/22, H3N2 CR/23, H3N2-K BA/25, and H3N2-K NY/25 were 46, 54, 57, 33, and 29 at baseline and increased to 115, 129, 147, 66, and 52 at peak immunogenicity, respectively (**Fig. 1C**), reflecting a 2.39- to 2.58-fold increase in HAI titers against the prior H1N1 and H3N2 strains but a lower 1.78- to 2.00-fold increase in HAI titers against the H3N2-K strains. Binding antibody responses were assessed by enzyme-linked immunosorbent assays (ELISA) and electrochemiluminescence assays (ECLA) against prior H1N1 and H3N2 strains also showed increases following vaccination (**Figs. S2, S3**).

Our data show that baseline antibody responses to two H3N2 subclade K variants were lower than to other H1N1 and H3N2 strains and that antibody responses following vaccination were also less robust to the H3N2 subclade K variants. These data indicate that the H3N2 subclade K strain partially evades both prior H3N2 immunity and the current inactivated influenza vaccine. The modest boost of antibody titers following immunization nevertheless suggests that the vaccine will still provide some benefit against H3N2 subclade K. However, the lower antibody titers to H3N2 subclade K compared with other recent influenza strains, both before and after vaccination, has important implications for the likely continued spread of this new influenza strain.

## Data Availability

All data produced in the present work are contained in the manuscript

## Author Contributions

L.W. and D.H.B. conceptualized the study. L.W., S.N., A.W.P., K.M., J.P., J.C., and L.O.B. performed the assays. R.B. and A.Y.C. led the clinical biorepository. K.F., R.R., E.M.S., V.S., H.V.B., H.M., and A.P. provided the H3N2 viruses. L.W. and D.H.B. wrote the manuscript with all co-authors.

## Acknowledgments

We thank BEI Resources for providing the H3N2/Baltimore/JH-335/2022 virus strain (NR-59460). We thank the study participants and the BIDMC Clinical Research Center and MesoScale Discovery for providing the ECLA kits. We thank Adolfo Gastre-Sastre, Kaijun Jiang, Zain Khalil, Yee Vue, Elgin Akin, Matthew Pinsley, Anne Werner, Erica Borducchi, Jinyan Liu, Joseph Nkolola, Ninaad Lasrado, Siddhesh Warke, Revant Singh, Nicole Berglund, Arunaa Ganesan, Bridget Wixted, Krishna Shah, Rae Filley, Valerie Perinnez, Amanda Michael, and Eleanor Nicholson for generous advice, assistance, and reagents.

## Supplementary Methods

### Study Population

We conducted a descriptive cohort study evaluating immune responses in 46 healthy adults who received the inactivated trivalent influenza vaccine in fall 2025 in Boston, MA. The specimen biorepository at Beth Israel Deaconess Medical Center (BIDMC) obtained peripheral blood samples from the study participants. The BIDMC institutional review board approved this study (2020P000361). All participants provided informed consent. Participants provided their vaccination, infection, and medical history as well as their self-reported race and ethnicity. Participants were excluded from the immunologic assays if they had a recent history of influenza infection within 2 weeks of enrollment or if they received immunosuppressive medications. Assay operators were blinded to participant infection and vaccination history. Participant demographics are shown in **Table 1**.

### Neutralization Assay

Neutralization titers of samples were assessed using the TCID_50_ reduction assay. Briefly, samples were serially diluted in 50 μL DMEM, 1% bovine albumin fraction, 1% Pen-Strep and incubated with 100 TCID_50_ of A/H1N1/Wisconsin/67/2021 (GISAID ID: EPI_ISL_19440421), A/H3N2/Baltimore/JH-335/2022 (GISAID ID: EPI_ISL_18143304), A/H3N2/Croatia/10136RV/2023 (GISAID ID: EPI_ISL_19296516), A/H3N2-K/Baltimore/JH-1365/2025 (GISAID ID: EPI_ISL_20281465), or A/H3N2-K/New York City/PX23710/2025 (GISAID ID: pending) in 50 μL for 1 hour at 37 °C in a 5% CO_2_ incubator. During the incubation, the MDCK-SIAT1 cell monolayer was treated with trypsin-EDTA, and the cells were resuspended in virus diluent. Subsequently, 1.5x10^4^ cells in 100 μL were added to the sample-virus mixture and incubated at 37 °C in a 5% CO_2_ incubator for 18-20 hours. The following day, the cell monolayer was fixed with 200 μL 80% cold acetone for 10 minutes after discarding the supernatant. The plates were dried and then blocked with 1% bovine serum albumin (BSA) in DPBS for 1 hour. The amount of the virus in each well was quantified by ELISA using anti-influenza A virus nucleoprotein (NP) monoclonal antibodies Clone A and Clone 3 at a 1:4000 dilution for each (Sigma, MAB8257; MAB8258) or anti-influenza B virus NP monoclonal antibodies at a 1:2000 dilution (Invitrogen, MA1-80712), and HRP-conjugated secondary antibodies at a 1:10000 dilution (VWR 95058-736) in 1% BSA. After washing the plates five times, 100 μL of SeraCare KPL TMB SureBlue start solution was added to each well. After 4 minutes, the reaction was halted by adding 100 μL of SeraCare KPL TMB stop solution per well. The neutralization antibody titer was defined as the reciprocal of the highest serum dilution that provided ≧ 50% inhibition of virus infectivity in absorbance was observed relative to the average of the virus control and cell control wells, determined using a four-parameter logistic curve fit in GraphPad Prism.

### Hemagglutination Inhibition (HAI) Assay

Serum samples were treated with receptor-destroying enzyme (RDE; Denka Seiken, Tokyo, Japan) by adding 3 volumes of RDE to 1 volume of sera and incubated at 37 °C for 18-20 h to remove nonspecific inhibitors of hemagglutination. After adding 6 volumes of saline solution to obtain a 1:10 starting dilution, serum samples were then heat-treated at 56 °C for 1 hour. Samples were 2-fold serially diluted in 25 ul of PBS in 96-well V-bottom microtiter plates and then incubated with 25 ul of 4 HA units of each virus for 30 mins at room temperature. The plate was incubated another 30 minutes at room temperature after which 50 ul of turkey red blood cells (Lampire Biological: 724940815ML) at 0.5% in PBS was added. HAI titer was calculated with the highest serum dilution that completely prevented agglutination.

### Enzyme-linked immunosorbent assay (ELISA)

Influenza HA-specific binding antibodies in serum were assessed by ELISA. 96-well plates were coated with 1 μg/mL of HA protein from A/H1N1/Wisconsin/67/2021 or A/H3N2/Darwin/6/2021 in Dulbecco phosphate-buffered saline (DPBS) and incubated at 4 °C overnight. After incubation, plates were washed once with wash buffer (0.05% Tween 20 in 1× DPBS) and blocked with 350 μL of casein block solution per well for 2 to 3 hours at room temperature. Following incubation, block solution was discarded and plates were blotted dry. Serial dilutions of heat-inactivated serum diluted in Casein block were added to wells, and plates were incubated for 1 hour at room temperature, prior to 3 more washes and a 1-hour incubation with a 1:4000 dilution of antihuman IgG horseradish peroxidase (HRP) (Invitrogen, ThermoFisher Scientific) at room temperature in the dark. Plates were washed 3 times, and 100 μL of SeraCare KPL TMB SureBlue Start solution was added to each well; plate development was halted by adding 100 μL of SeraCare KPL TMB Stop solution per well. The absorbance at 450 nm was recorded with a VersaMax microplate reader (Molecular Devices). For each sample, the ELISA end point titer was calculated using a 4-parameter logistic curve fit to calculate the reciprocal serum dilution that yields a corrected absorbance value (450 nm-650 nm) of 0.2. Interpolated end point titers were reported.

### Electrochemiluminescence assay (ECLA)

ECLA plates [MesoScale Discovery (MSD); V-PLEX Respiratory catalog no. K15735U (panel 7)] were designed and produced with up to 9 antigen spots in each well to detect IgG antibodies to a variety of important respiratory pathogens. The antigens included the HA proteins from A/H1N1/Wisconsin/67/2022 or A/H3N2/Massachusetts/18/2022. The plates were blocked with 50 μl of blocker A (1% bovine serum albumin in distilled water) solution for at least 30 min at room temperature with shaking at 700 rpm with a digital microplate shaker. During blocking, serum samples were diluted to 1:5000 in Diluent 100. The calibrator curve was prepared by diluting the calibrator mixture from MSD 1:9 in Diluent 100 and then preparing a seven-step fourfold dilution series and a blank containing only Diluent 100. The plates were then washed three times with 150 μl of wash buffer [0.5% Tween 20 in 1× phosphate-buffered saline (PBS)] and blotted dry, and 50 μl of the diluted samples and calibration curve was added in duplicate to the plates and set to shake at 700 rpm at room temperature for at least 2 hours. The plates were again washed three times, and 50 μl of SULFO-TAG anti-human IgG detection antibody diluted to 1× in Diluent 100 was added to each well. Samples were incubated with shaking at 700 rpm at room temperature for at least 1 hour. Plates were then washed three times, and 150 μl of the MSD GOLD Read Buffer B was added to each well. The plates were read immediately after using a MESO QuickPlex SQ 120 machine. MSD titers for each sample were reported as relative light units, which were calculated using the calibrator.

## Supplementary Figure Legends

**Supplemental Figure 1.**
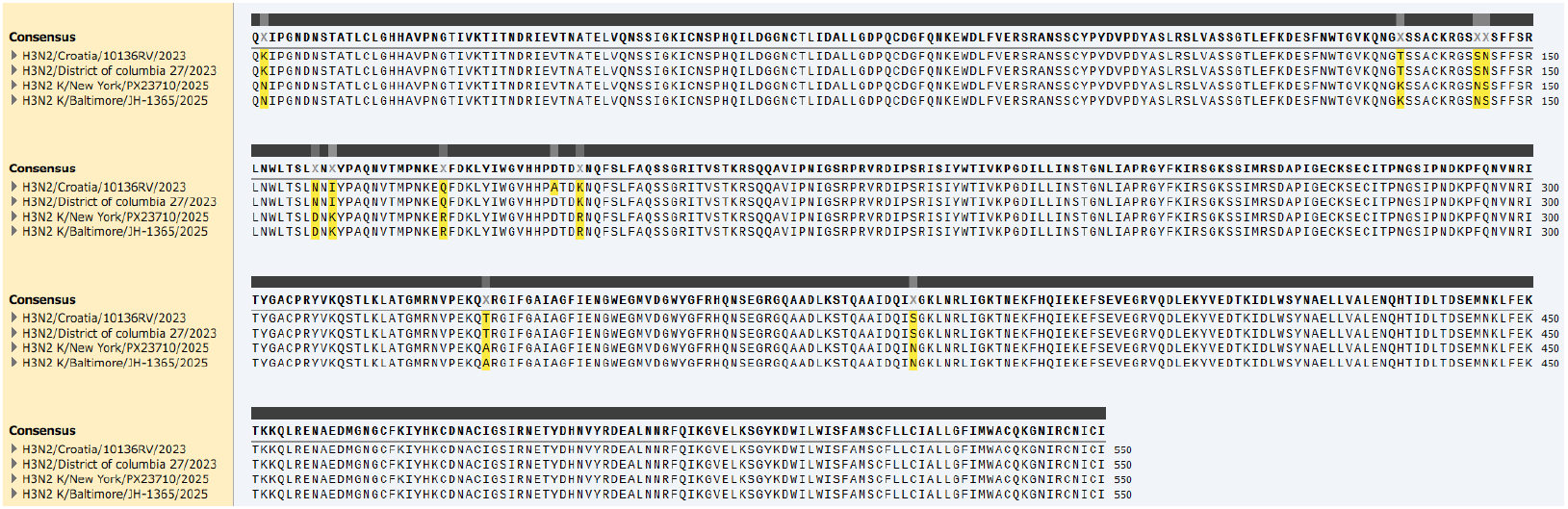
Sequence alignment of HA proteins from the H3N2 vaccine strains A/H3N2/Croatia/10136RV/2023 and A/H3N2/District of Columbia/27/2023 compared with the H3N2 subclade K strains A/H3N2-K/New York City/PX23710/2025 and A/H3N2-K/Baltimore/JH-1365/2025. There are 11 amino acid differences between the subclade K strains and the A/H3N2/Croatia/10136RV/2023 vaccine strain and 10 amino acid differences between the subclade K strains and the A/H3N2/District of Columbia/27/2023 vaccine strain.

**Supplemental Figure 2.**
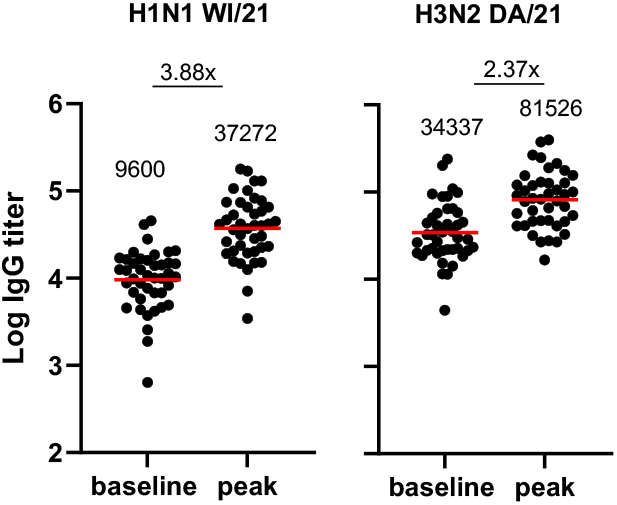
IgG binding antibody responses by ELISA before (baseline) and after (peak) vaccination to HA antigens from A/H1N1/Wisconsin/67/2021 (H1N1 WI/21) or A/H3N2/Darwin/6/2021 (H3N2 DA/21). Red bars indicate geometric mean values.

**Supplemental Figure 3.**
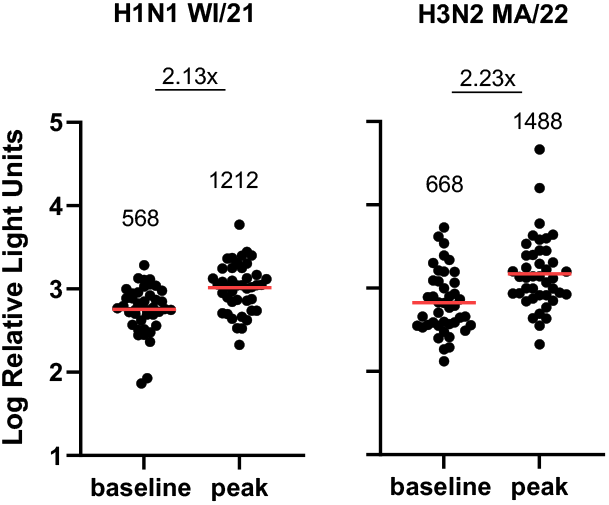
IgG binding antibody responses by ECLA before (baseline) and after (peak) vaccination to HA antigens from A/H1N1/Wisconsin/67/2022 (H1N1 WI/21) or A/H3N2/Massachusetts/18/2022 (H3N2 MA/22). Red bars indicate geometric mean values.

